# Disparity in the quality of COVID-19 data reporting across India

**DOI:** 10.1101/2020.07.19.20157248

**Authors:** Varun Vasudevan, Abeynaya Gnanasekaran, Varsha Sankar, Siddarth A. Vasudevan, James Zou

**Affiliations:** Institute for Computational & Mathematical Engineering, Stanford University; Independent Researcher, USA; Independent Researcher, Switzerland; Department of Biomedical Data Science, Stanford University

## Abstract

**Background:** Transparent and accessible reporting of COVID-19 data is critical for public health efforts. Each state and union territory (UT) of India has its own mechanism for reporting COVID-19 data, and the quality of their reporting has not been systematically evaluated. We present a comprehensive assessment of the quality of COVID-19 data reporting done by the Indian state and union territory governments. This assessment informs the public health efforts in India and serves as a guideline for pandemic data reporting by other governments.

**Methods:** We designed a semi-quantitative framework to assess the quality of COVID-19 data reporting done by the states and union territories of India. This framework is based on 45 indicators that capture four key aspects of public health data reporting – availability, accessibility, granularity, and privacy. We then used this framework to calculate a *COVID-19 Data Reporting Score* (CDRS, ranging from 0 to 1) for 29 states^i^ based on the quality of COVID-19 data reporting done by the state during the two-week period from 19 May to 1 June, 2020. States that reported less than 10 total confirmed cases as of May 18, were excluded from the study.

**Findings:** Our results indicate a strong disparity in the quality of COVID-19 data reporting done by the state governments in India. CDRS varies from 0.61 (good) in Karnataka to 0.0 (poor) in Bihar and Uttar Pradesh, with a median value of 0.26. Only ten states provide a visual representation of the trend in COVID-19 data. Ten states do not report any data stratified by age, gender, comorbidities or districts. In addition, we identify that Punjab and Chandigarh compromised the privacy of individuals under quarantine by releasing their personally identifiable information on the official websites. Across the states, the CDRS is positively associated with the state’s sustainable development index for good health and well-being (Pearson correlation: *r* = 0.630, *p* = 0.0003).

**Interpretation:** The disparity in CDRS across states highlights three important findings at the national, state, and individual level. At the national level, it shows the lack of a unified framework for reporting COVID-19 data in India, and highlights the need for a central agency to monitor or audit the quality of data reporting done by the states. Without a unified framework, it is difficult to aggregate the data from different states, gain insights from them, and coordinate an effective nationwide response to the pandemic. Moreover, it reflects the inadequacy in coordination or sharing of resources among the states in India. Coordination among states is particularly important as more people start moving across states in the coming months. The disparate reporting score also reflects inequality in individual access to public health information and privacy protection based on the state of residence.

**Funding:** J.Z. is supported by NSF CCF 1763191, NIH R21 MD012867-01, NIH P30AG059307, NIH U01MH098953 and grants from the Silicon Valley Foundation and the Chan-Zuckerberg Initiative.

## 1 Introduction

India reported its first case of COVID-19 in the state of Kerala on January 30, 2020. Since then the disease has been reported in several other states and union territories (UTs) of India. As of July 18, 2020, the Ministry of Health and Family Welfare (MoHFW) of India reported over a million COVID-19 confirmed cases and over twenty-six thousand COVID-19 deaths in the country.^1^ India is a developing nation and has the second largest population in the world. India is also a democracy with 28 states and 8 union territories. Therefore, coordinating an effective response to the pandemic, across all the regions, presents a unique and unprecedented challenge to India.

Both the central and state governments in India have introduced multiple measures and interventions for the containment of COVID-19.^2^ It is well known that public adherence to these measures and interventions is essential for managing the pandemic.^3^ In order to keep the public informed about the ongoing situation, the states in India have been reporting COVID-19 data. Reporting relevant data in a timely, transparent, and accessible manner is crucial during a pandemic. The advantages of such a timely reporting are atleast two-fold. First, it fosters trust between the government and the public and, thereby ensures public cooperation. Second, it enables the scientific community to rapidly and continually study the reported data to gain insights and propose better containment measures and policies. A schematic of a good data reporting system is shown in Appendix A.

The content and format of the COVID-19 data reports vary substantially from state to state. Figure 1 shows how total (cumulative) numbers are reported by three different states in India. Notice how Assam and Gujarat report just the total numbers, whereas Kerala reports the numbers and their trend graphics. In addition to reporting the numbers, providing trend graphics is essential because it concisely represents the data, and makes it more interpretable and accessible to the general public. The variance in reporting across states raises two key questions. First, what is the minimal data that the public needs to know to understand the gravity of the situation and cooperate with the government? Second, how different is the quality of data reporting from one state to another?

**Figure 1:**
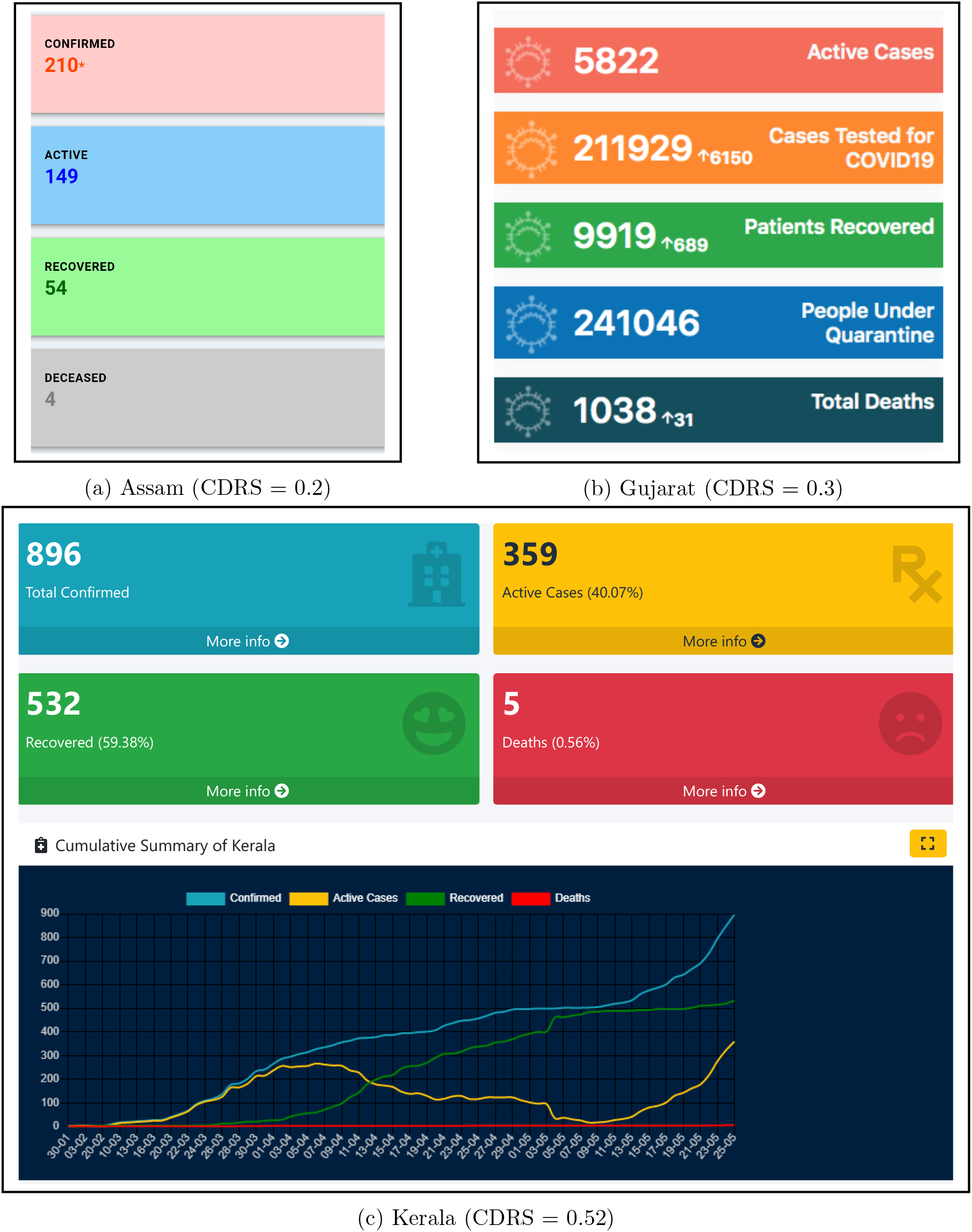
Screenshots from the government COVID-19 web pages of Assam, Gujarat, and Kerala displaying cumulative data. Kerala is the only state among the three to provide both a textual summary and trend graphics, and it has the second highest COVID-19 Data Reporting Score (CDRS).

Contributions: In this paper, we answer the two aforementioned questions by developing a systematic framework to evaluate the quality of COVID-19 data reporting. We then use it to assess the quality of reporting done by the states in India. Based on our framework we provide a minimal template that the states can use for daily COVID-19 data reporting (given in Appendix B). We also present our findings on an interactive dashboard (web link) that’s easily accessible.

### 1.1 Related work

Quality of data reporting: A leading Indian English newspaper, The Hindu, recently published an article showing variance in COVID-19 data reporting across the states in India.^4^ This article reflects the general public sentiment in seeking high quality data. However, their analysis has atleast three main limitations. First, it provides only a high-level summary of the variance in data reporting and is limited to 21 states. Second, the article focuses only on data reported in the health bulletins, whereas we inspected data reported in all formats on government websites. Third, they don’t provide a quantitative analysis. Janiaud and Goodman developed less granular metrics to assess reporting quality in the U.S. states.^5^

Data quality: Data quality is a multidimensional concept with dimensions such as, accuracy, accessibility, completeness, interpretability, relevancy, and timeliness.^6^ There are frameworks for data quality assessment that are motivated by what data quality means to the consumers of data.^7,8^ Although there is an overlap between quality of data and quality of data reporting, they are not quite the same. Accuracy is a crucial aspect in data quality. However, while measuring the quality of data reporting, the emphasis is not on the accuracy of data, instead it is on the presence or absence of a piece of information and the format in which it is reported.

Data visualization: Visualization is critical for understanding data. Excellent statistical graphics communicate complex ideas with clarity, precision, and efficiency.^9^ The best practices in creating statistical graphics are discussed extensively in the books by Cleveland and Tufte.^9,10^ There is also rich literature on developing effective real-world dashboards,^11^ and interactive visualization for the web.^12^ We check for the presence of visuals in the form of trend graphics while assessing the quality of data reporting. However, we do not assess the attributes of a graphic such as shape (length to height ratio), line weight, choice of colors, font size of text, and whether the graphic is interactive or not.

Crowdsource initiative for COVID-19 data: covid19india.org^ii^ is a volunteer driven crowd-sourced tracker for COVID-19 cases in India. They collect and curate COVID-19 data from all across India, from a variety of sources, including but not limited to state government websites.^13^ The curated data is reported on their website in the form of tables, trend graphics, and color-filled maps. They also provide an option to download data. The covid19india team has an active page on Twitter with more than 100 thousand followers. Based on the number of followers and the kind of questions^iii^ they ask, it is evident that people are seeking granular COVID-19 data on a daily basis. This crowdsource initiative is a commendable example for public participation during a crisis. Nevertheless, it is not sufficient, and does not replace the need for clear and consistent government official reporting for the following reasons. The initiative is volunteer driven and hence accountability is not guaranteed in the event of an error or lapse in reporting. Moreover, their sources for data include social media, which are noisy.

### 2 Methods

We developed a set of metrics to score the quality of “COVID-19 data reporting” done by the states in India. These metrics are shown in column 2 of Table 1. The metrics are further grouped into four categories: availability, accessibility, granularity, and privacy, as shown in column 1 of Table 1. Using these metrics, we examine the quality of reporting for five items relevant to COVID-19.They are confirmed, deaths, recovered, quarantine and intensive care unit (ICU) cases. These are called as report items and appear as column headers in the scoring table. The report items represent five possible stages through which an individual can go through during a pandemic. For example, an individual could move from the stage of being under quarantine, to being a confirmed case, and from there could recover in a couple of weeks, or if the situation worsens, could move to ICU. At the time of this study, all confirmed COVID patients in India were hospitalized and treated in one of the following facilities: COVID Care Centers, Dedicated COVID Health Centers or Dedicated COVID Hospitals.^14^ Each “metric report item” pair is a data reporting quality indicator (variable). Overall there are 45 indicators in our framework across the four scoring categories. It is important to note that neither the list of metrics, nor the list of report items used in our scoring table are exhaustive. It is a representative minimal set.

**Table 1:**
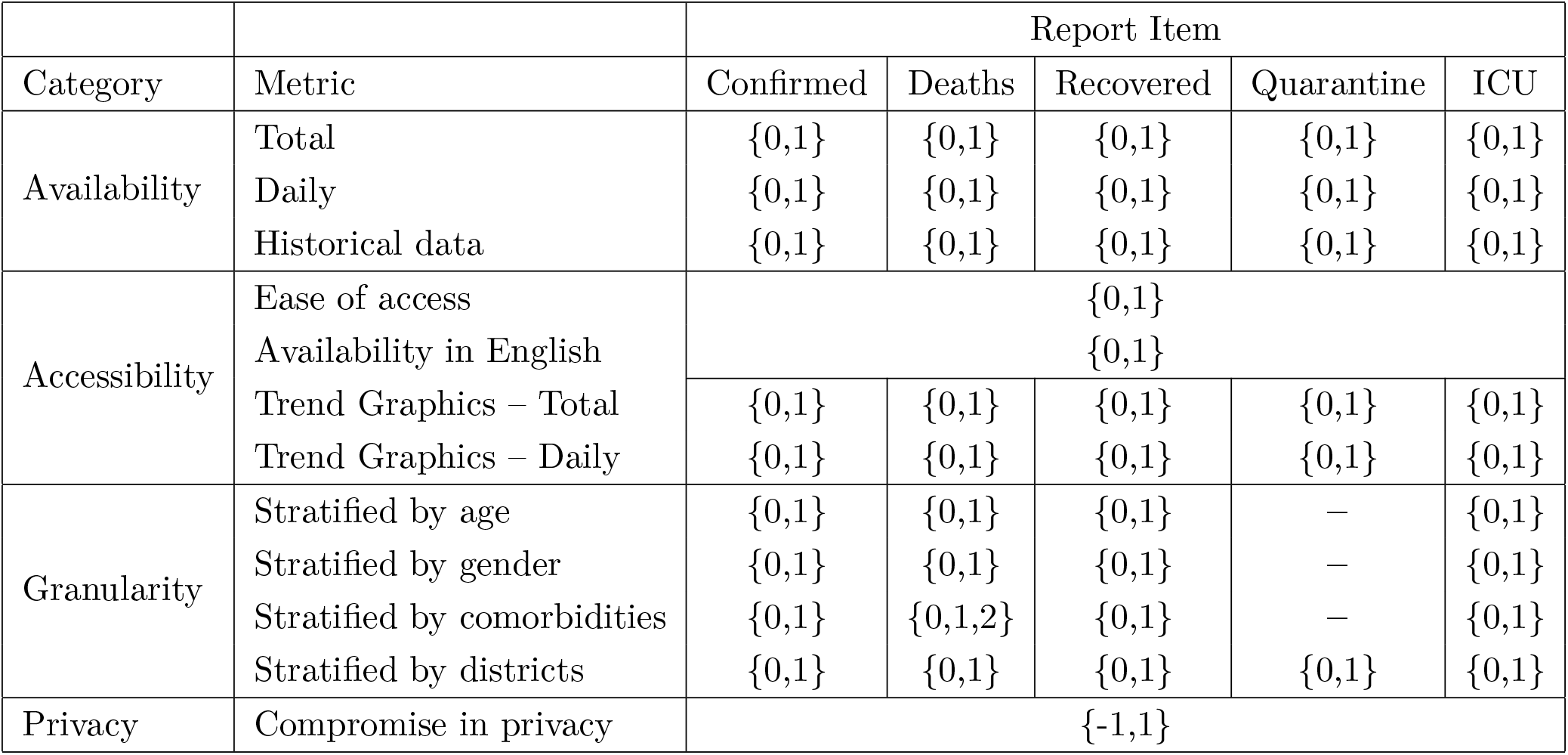
CDRS Scoring metric table. This table is filled for each state by inspecting the COVID-19 data reported by that state. The entry within a cell in the table lists all the possible values with which that cell can be filled. Broadly, a 0 represents an unreported item, and a 1 represents a reported item.

We define the report items as follows.

1. (Laboratory) Confirmed: refers to individuals who tested positive for COVID-19.
2. Deaths: refers to individuals who passed away while being COVID-19 positive.
3. Recovered: refers to individuals who recovered from COVID-19.
4. Quarantine: refers to individuals who are under quarantine either at home or specific government facilities. The definition of who should be quarantined and for how long has evolved during the course of pandemic in India.
5. ICU: refers to COVID-19 positive individuals who are under treatment in an ICU.

Our metrics and report items are in line with World Health Organization’s (WHO) COVID-19 surveillance guidance,^15^ and the questions posed in the paper, “Transparency during public health emergencies: from rhetoric to reality“.^3^ The latter identifies three YES/NO questions to help in deciding whether or not to release a piece of information related to a public health emergency. These questions seek to understand the role of a piece of information in: (i) reducing the spread of a disease, (ii) emergency management decision making process, and (iii) compromising privacy or stigmatization of specific groups of people or both.

### 2.1 Scoring categories

In this section we give an overview of the four scoring categories. For more information on each scoring metric listed in Table 1 refer to Appendix D.

1. Availability of data: During a pandemic, few generic questions that people seek to answer are: “How are we doing?“, “How do we know how we are doing?“, “How long will this last?“, “How do the numbers from today compare with yesterday’s?“, “How many people have tested positive so far“, and so on. With such questions in mind we measure the availability of data by checking if the total, daily, and historical data are available for each report item.
2. Accessibility of data: Data should not only be available, it should also be easily accessible. We measure the accessibility of data based on ease of access, availability in English, and the presence of trend graphics. Ease of access refers to the ease of getting to the web page where data is reported. Research has shown that trend graphics are superior than tables for identifying and displaying trends.^16^ A good visual concisely represents the data and makes it more interpretable and accessible to the general public. Therefore, to measure accessibility we also check, if a graphic of total and daily are available for each report item.
3. Granularity of data: Granularity refers to the stratification of the total number for each report item. We check if the total is stratified by age, gender, comorbidities, and districts. Recent studies have shown the role of age, gender, and comorbidities in influencing the outcome of a COVID-19 positive individual.^17–19^ As per the Indian Council of Medical Research (ICMR) specimen referral form for COVID-19, data on age, gender, district, and pre-existing medical conditions are collected for each person being tested.^20^ Therefore, aggregating and then stratifying that information should be straightforward. At a higher level stratified information is useful in the following ways. (i) District level data keeps the public informed about the gravity of situation in their neighborhood. (ii) People can self-identify how susceptible they are to get infected and hence take the necessary precautions. For example, granular data can answer questions of the kind, “I’m 65 and healthy, should I be worried?” (iii) Scientific community can study the effect of factors like age, gender, and comorbidities on contracting the disease, its progression, and the outcome.
4. Privacy of data: Data released by the government should include only the minimum information necessary to conduct public health activities.^21^ It should not contain any personally identifiable information. Violating privacy by releasing personally identifiable information can have the following consequences. (i) It can discourage people from cooperating with the government, thereby hurting public health rather than helping. (ii) Women can be victims of harassment calls when their phone number is released. A study by Truecaller shows that, in general, 8 out of 10 women in India receive harassment and nuisance calls.^22^ Releasing phone numbers can further amplify the general trend. (iii) Discrimination and stigmatization of specific groups of people.^23–25^

### 2.2 Scoring data curation

We collected the data reported by the states during the two week period from May 19, 2020 to June 1, 2020 for assessing the quality of “COVID-19 data reporting“. Hereafter, this collected data is referred to as the *scoring data* and the two week period is referred to as the *scoring period*. By May 18, India was already under lockdown for more than 50 days. This is sufficient time for state governments to develop a good data reporting system. The fact that India had reported 96 thousand confirmed cases by then made it all the more important to warrant a high quality data reporting system. Therefore, our choice of scoring period is reasonable and the scoring data collected during this period captures a quasi-steady state for reporting. States that reported less than 10 total confirmed cases as of May 18, were excluded from the study. The excluded states were: Arunachal Pradesh, Dadra and Nagar Haveli and Daman and Diu, Lakshadweep, Manipur, Mizoram, Nagaland and Sikkim. After the exclusion we were left with 29 states for assessment. In each of the 29 states, the first case^iv^ was reported atleast 30 days prior to May 19. The authors applied the scoring criteria in Table 1 to each state and reached a consensus on the score for each state (details of the consensus scoring process is described in Appendix E).

### 2.3 Score calculation

Scoring data was curated for 29 states using the procedure described in the previous section. For each of these states we calculate four categorical scores — availability, accessibility, granularity, and privacy, and an overall score, which is referred to as the COVID-19 Data Reporting Score (CDRS). CDRS is the normalized sum of these four categorical scores and ranges from 0 (lowest quality) to 1 (highest quality). Categorical scores for a state are calculated by summing the entries corresponding to that category in the scoring table. All reported items are weighted equally while calculating the scores.

CDRS and the normalized categorical scores for the 29 states are available in Appendix F. The normalized scores for availability, accessibility, and granularity range from 0 (lowest value) to 1 (highest value). The normalized privacy score is 0.5 when there is no violation of privacy and −0.5 otherwise. Privacy score is not applicable for states that do not report any data. For all the score calculations, normalization was adjusted to account for not applicable (’NA’) entries in the scoring metric table (see Appendix E).

## 3 Results

As described in the previous section, a COVID-19 Data Reporting Score (CDRS), and four normalized categorical scores were calculated for 29 states in India.

There is a strong disparity in the quality of COVID-19 data reporting done by the different states. The five number summary of CDRS is, min = 0.0, first quartile = 0.2, median = 0.26, third quartile = 0.41, and maximum = 0.61. The disparity can be clearly seen in Figure 2, which shows the CDRS for the different states, both as a color-filled map and as a dot plot. The boundary information for regions in India was obtained as shapefiles from Datameet (http://projects.datameet.org/maps/). Visuals for the normalized availability, accessibility, granularity, and privacy scores are available in Appendix F.

**Figure 2:**
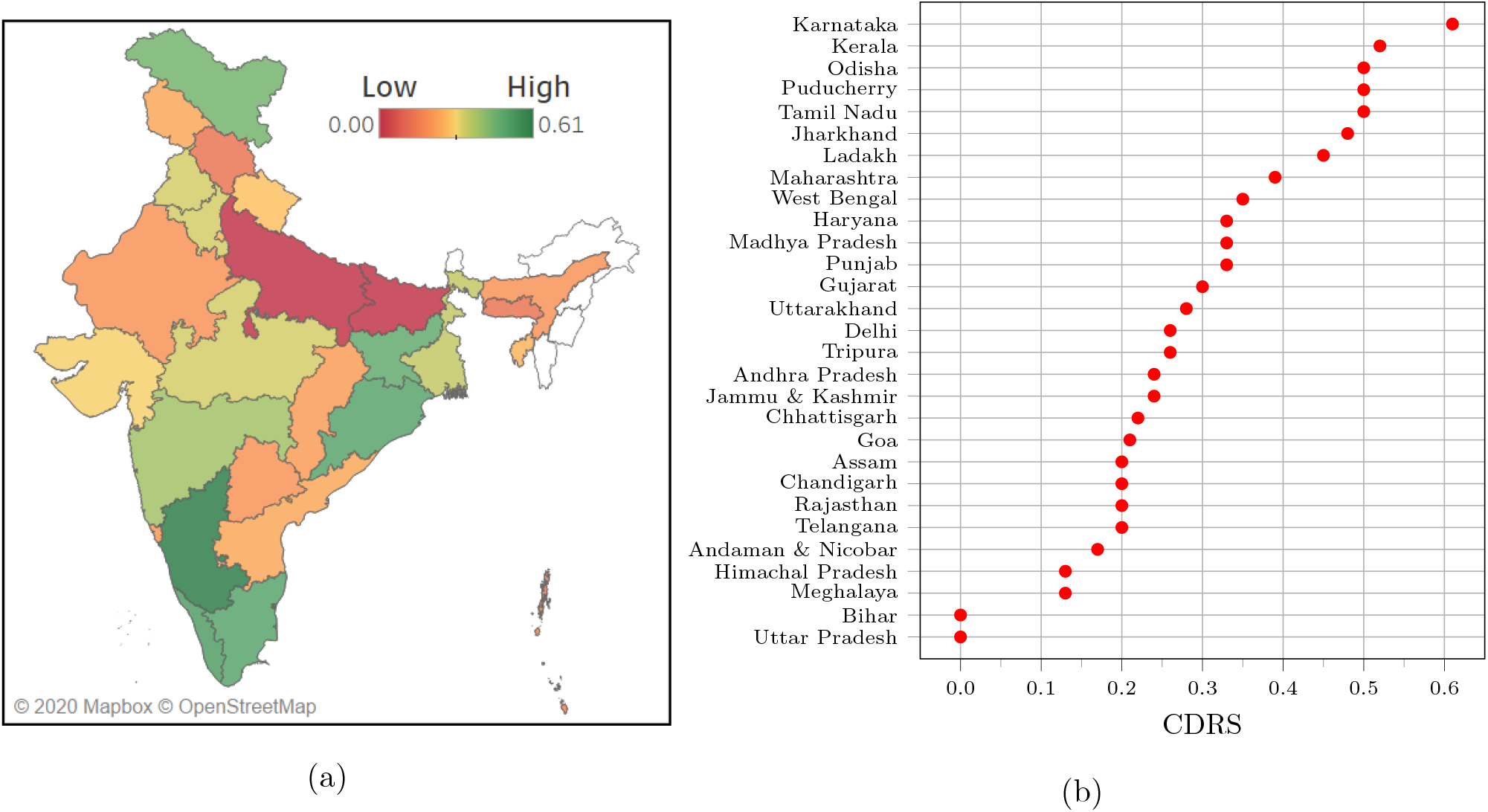
(a) Map showing CDRS across the states and UTs in India. States and UTs that were excluded from our study are filled with a white color. The map clearly shows the geographical disparity in COVID-19 data reporting in India. The map was generated using Tableau Desktop software version 2020.2.1 and the boundary information for regions in India was obtained as shape-files from Datameet (http://projects.datameet.org/maps/). (b) A dot plot showing the spread of CDRS values. On the y-axis, states are sorted in the decreasing order of CDRS.

The best data reporting is done by Karnataka (0.61), Kerala (0.52), Odisha (0.50), Puducherry (0.50), and Tamil Nadu (0.50). On the other hand, Uttar Pradesh (0.0), Bihar (0.0), Meghalaya (0.13), Himachal Pradesh (0.13), and Andaman and Nicobar Islands (0.17) rank at the bottom. For details on the number of states that report a specific information, refer to Figure 3.

**Figure 3:**
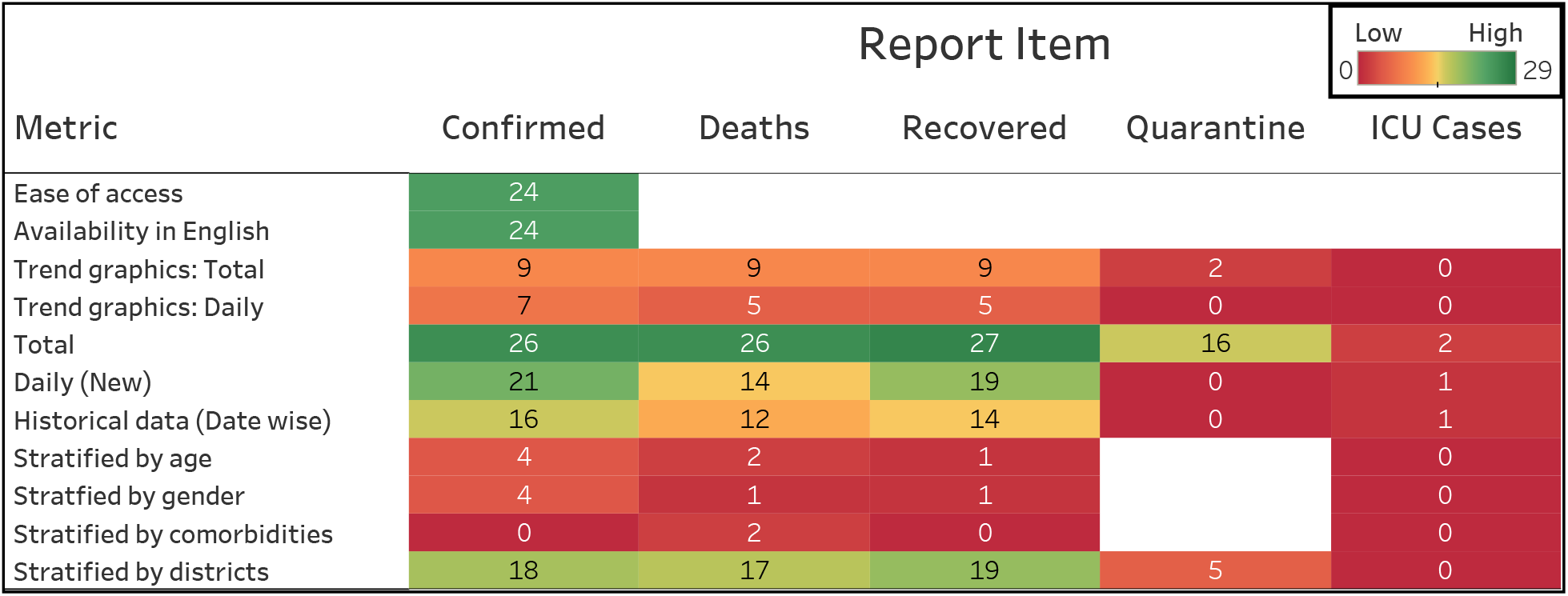
Table shows the number of states reporting an item out of twenty-nine states. Twenty-six out of the twenty-nine states report the total number of COVID-19 confirmed cases and deaths, and twenty-seven states report the number of recovered individuals. Only a handful of the states publish cumulative data stratified by age, gender, and comorbidities.

As seen in the Figure 2, CDRS varies from 0.61 in Karnataka to 0.0 in Bihar and Uttar Pradesh. Figure 4 shows few screenshots from the bulletins released by Karnataka on May 31, 2020. Karnataka’s COVID-19 page is linked from the state government’s website. The state releases a health bulletin and a state war room bulletin everyday, and also maintains a dashboard. The bulletins are available in English, and provides information on the total confirmed, deaths, recovered, quarantined, and active ICU cases. The bulletins also report some daily (new) data, and some data stratified by age, gender, and districts. In addition, the demographics and comorbidity data are reported for each deceased person. Trend graphics are available either through the bulletins and/or through the dashboard. Bihar and Uttar Pradesh do not publish any data on their government or health department website. Hence their CDRS is 0.

**Figure 4:**
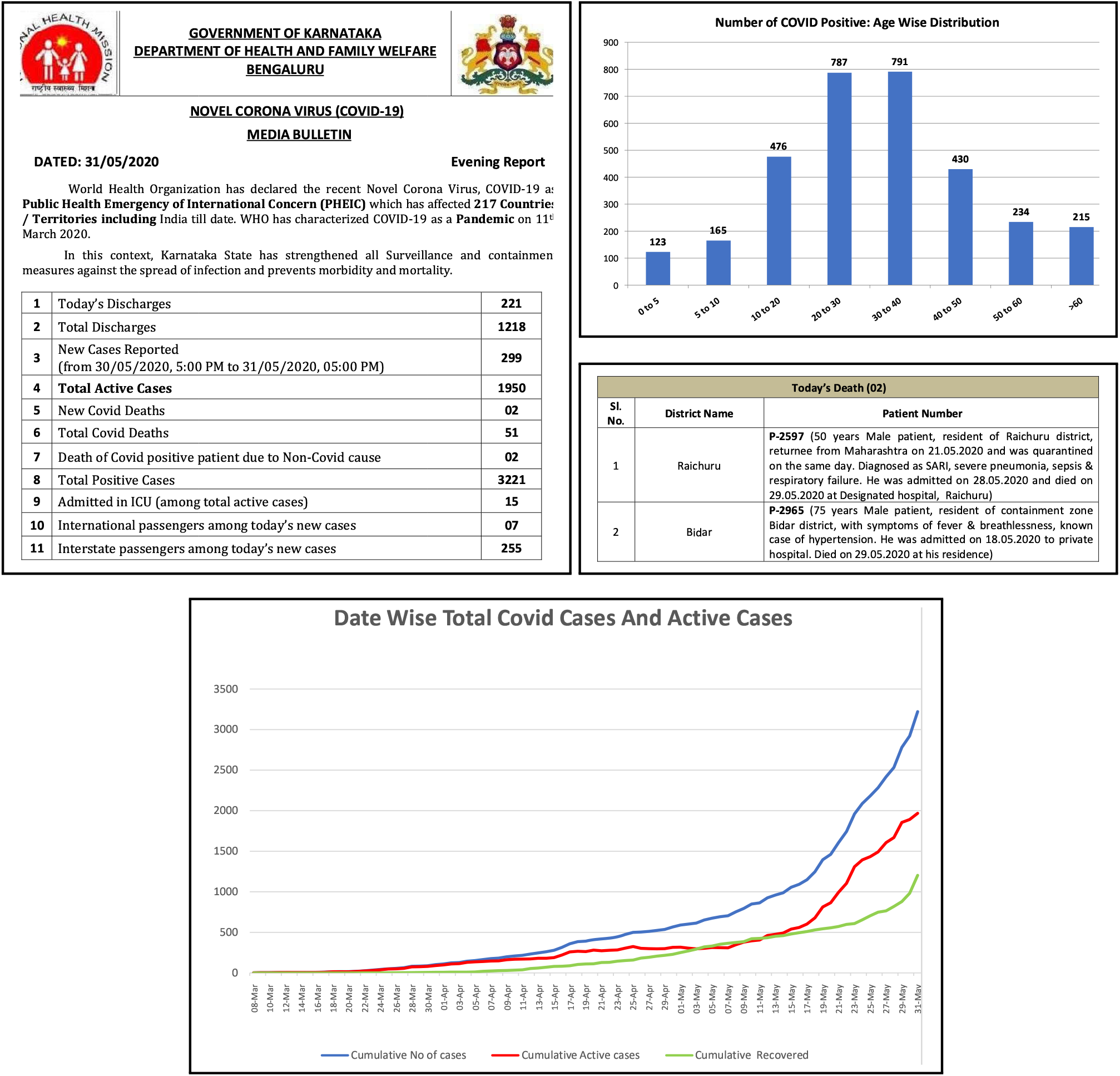
Screenshots from the bulletins released by Karnataka government on May 31, 2020 as examples of high quality COVID-19 Data Processing.

Karnataka and Punjab score the highest in availability. Both these states report the daily and total numbers for confirmed, deceased and recovered cases. They also report COVID-19 cases in Intensive Care Units (ICUs). Historical data is available for both the states in the form of daily bulletins. Among the states that report data, Assam, Himachal Pradesh, and Meghalaya score the lowest for availability. This is because they report only the total count for confirmed, deceased, and recovered. A screenshot of the data reported by Assam is shown in Figure 1 along with its CDRS.

COVID-19 data can be accessed from the state’s official websites for 83% of the states. Only 10 states make the data more accessible by providing a visual representation of the trend. Karnataka and Kerala score the highest (0.75) in the accessibility category. These states provide trend graphics for both total and daily data, for the confirmed, deceased, and recovered cases. The bottom row in Figure 1 shows the screenshot of a trend graphic displayed on Kerala government’s COVID-19 dashboard.

In general, the worst categorical scores are for granularity. Even Jharkhand, the top state in this category, scored only a 0.50, while the median normalized granularity score is 0.17. For more details on the granular data published by Jharkhand refer to Appendix G. Karnataka and Tamil Nadu are the only states to provide details of death (including comorbidity information) for each deceased person. The following states do not report any data stratified by age, gender, comorbidities, or districts: Andaman and Nicobar Islands, Andhra Pradesh, Bihar, Chandigarh, Delhi, Goa, Himachal Pradesh, Meghalaya, Telangana and Uttar Pradesh.

Among the 29 states that were assessed 27 reported some data. Privacy doesn’t apply to states that do not report any data. Among the 27 states that report some data, all of them except Chandigarh and Punjab, report de-identified information and do not violate the privacy of the people residing in their state. Chandigarh has released name and residential address of people under home quarantine. Punjab has released name, gender, age, and mobile number of persons inbound to the state from New Delhi on May 10, 2020. Figure 5 shows screenshots from the documents published in the government websites of Punjab and Chandigarh that contain personally identifiable information.

**Figure 5:**
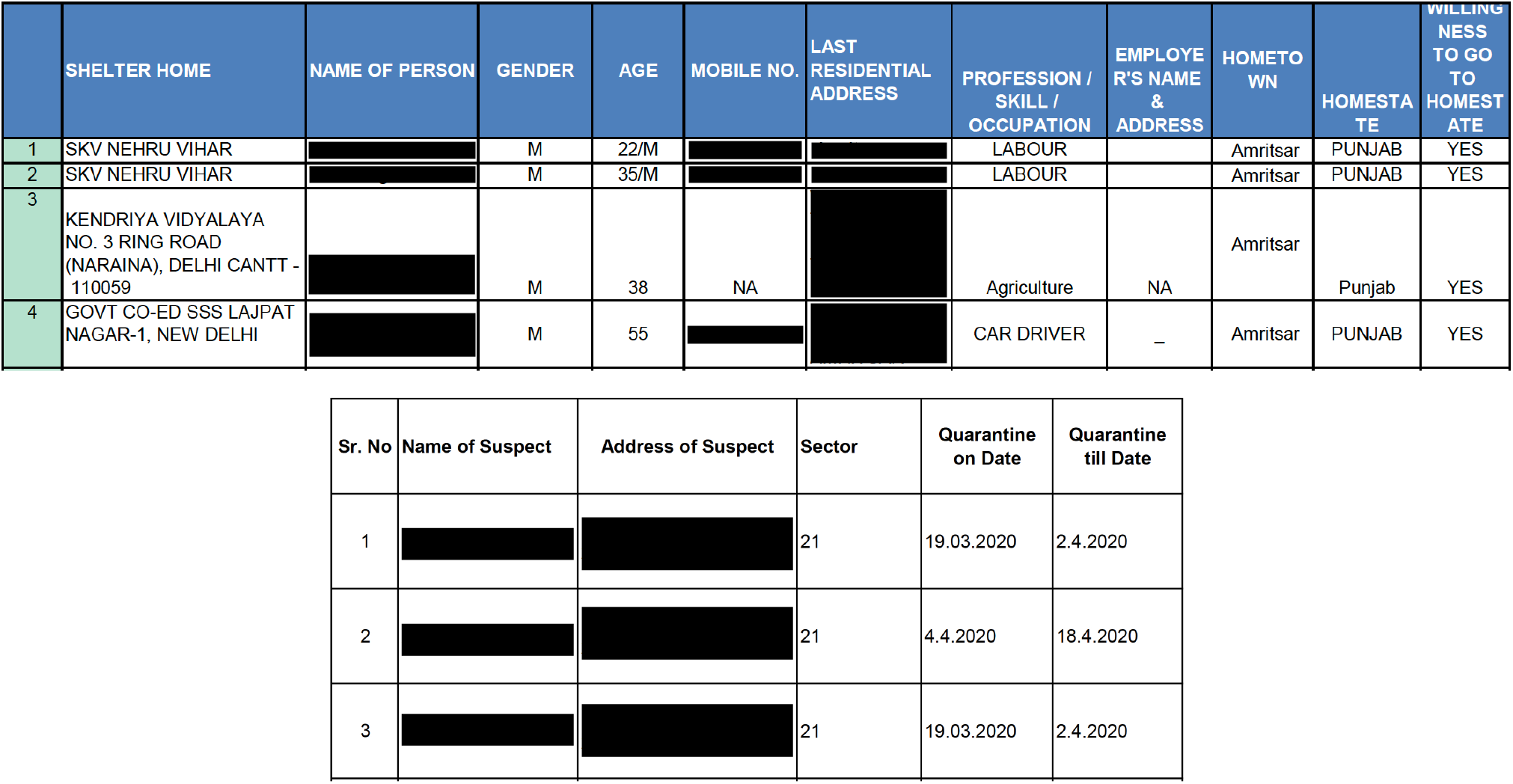
Screenshots of the documents published by Punjab (top row) and Chandigarh (bottom row) that contain individually identifiable information of people under quarantine. The individual’s name and address were reported on the official website and we blacked it out here.

We assessed the correlation between CDRS and the Sustainable Development Goal India Index for Good Health and Well-Being, abbreviated as SDG3-II. A positive correlation was observed between CDRS and SDG3-II (Pearson correlation: *r* = 0.630, *p* = 0.0003; and Spearman’s rank correlation: *r* = 0.578, *p* = 0.001). The positive correlation suggests that governments which are making more progress toward the “sustainable development goal of good health and well-being” also tend to have better COVID-19 data reporting. For more details on SDG3-II, refer to section 4.

## 4 Discussion

We assessed the quality of COVID-19 data reporting done by the states of India, during the two-week period from 19 May to 1 June, 2020. A total of 29 states were assessed during this time period. In each of the 29 states, the first case was reported atleast 30 days prior to May 19. In other words, each of these states had atleast a month’s time before the assessment to build a high-quality data reporting system. Our results and conclusions should be viewed and interpreted in light of this time frame.

The disparity in CDRS across the states highlights three important findings at the national, state, and individual level, respectively. First, it shows the lack of a unified framework for reporting COVID-19 data in India, and the need for a central agency to monitor or audit the quality of data reporting done by the states. Without a unified framework, it is difficult to aggregate the data from different states, gain insights from them, and coordinate an effective nationwide response to the pandemic. Not just that, unified high-quality data reporting also signifies transparency and hence increases public trust in the government. Containment becomes easier when the public is well-informed.

Second, it reflects the inadequacy in coordination or sharing of resources among the states in India. Coordination among states is particularly important as more people start moving across the states in the coming months. While it might not be possible for all the states to setup a high-quality dashboard in a short time, they can nevertheless, seek help and learn from the best data reporting practices followed by the other states.

Third, the disparate reporting score also reflects inequality in individual access to public health information and privacy protection based on the state of residence. The inequality highlights that the state-level efforts do not align with the central government’s vision of treating public health data as a public good, within the legal framework of data privacy, as described in the 2018–19 economic survey of India.^26^

States with high CDRS: Even Karnataka, the state with highest CDRS, has further scope for improvement, as its score is only 0.61. The top 5 states in CDRS provide a dashboard that shows the trend of COVID-19 data graphically. Also, they all provide district wise stratification of the total confirmed, recovered, and deaths due to COVID-19. However, not all of them stratify the data according to age, gender, and comorbidities, the factors that are known to have a correlation with the COVID-19 fatality rate.^17–19^

States with low CDRS: The states of Uttar Pradesh and Bihar do not publish any COVID-19 data on their government or health department website. However, Bihar seems to release some data on Twitter. See Appendix H for more details on that. We did not consider data reported on social media for multiple reasons. First, there are multiple social media platforms. Expecting people to be on the right platform and following the right person to obtain relevant public health information is unreasonable. Second, relevant information can easily get lost amid several posts. Third, obtaining historical data by scrolling through the feed is practically impossible. Himachal Pradesh, Meghalaya, and Andaman & Nicobar Islands, are the other three regions that rank in bottom 5 for CDRS. Each of them report just the total count for few report items. Daily count, trend graphics and granular data are not reported.

Privacy: We cannot stress enough about the importance of respecting the privacy of all citizens. One might argue that providing residential address of people under home quarantine is helpful to identify areas to avoid in a locality. However, the same information can be conveyed using hotspot maps that can be generated using geomasking techniques to protect privacy.^27^

CDRS and SDG3 India Index (SDG3-II): Sustainable Development Goals (SDGs) are a set of 17 global goals to achieve by 2030. These were set by the United Nations in 2015.^28^ The SDG India Index 2019–2020, developed by NITI Aayog, is a framework to measure the progress of states based on their performance across SDGs.^29^ The framework was developed using 100 indicators across 54 SDG targets. SDG3-II measures the performance of states on the third SDG, which is, Good Health and Well-Being for all. The value for SDG3-II ranges from 0–100, where 100 implies that the state has achieved the target set for the year 2030. Figure A.4 of Appendix I shows a scatter plot that displays the relationship between CDRS and SDG3-II. A positive correlation between CDRS and SDG3-II suggests that governments which are making more progress toward the “sustainable development goal of good health and well-being” also tend to have better COVID-19 data reporting. The indicators used to calculate SDG3-II are listed in Appendix I.

CDRS and total confirmed COVID-19 cases: According to MoHFW as of May 18, 2020, the total number of confirmed cases in India were about ninety-six thousand. The top ten states when sorted according to the number of confirmed cases contributed to a staggering 91% of the total confirmed. These ten states are shown in Figure A.5 of Appendix K above the horizontal dashed line. Tamil Nadu is the only state among those ten states with a CDRS in the 75th percentile. This suggests that states with the highest number of cases also tend to have poor COVID-19 data reporting, which could further exacerbate the pandemic challenges.

Overall, our scoring framework and CDRS together helps in identifying the differences in the quality of COVID-19 data reporting across India. In addition to revealing the disparity in the quality of reporting, CDRS also highlights that there is tremendous scope for all states to improve. The categorical scores enable states to identify their strengths and weaknesses. In each category, states can learn from their peers and improve their quality of reporting. States that score high in a category can serve as role models to the other states.

## 5 Limitations and future work

Some of the limitations of our study are as follows. (i) We did not include the reporting of testing data in our framework. This is because the degree of relevance of testing data in understanding the course of pandemic depends on whether testing was done on a scientific random sampling basis or not. (ii) Some states in India have developed mobile applications for COVID-19. We were unable to download and install them due to geographical restrictions. Therefore, our study doesn’t consider data that states might be reporting through these mobile applications. (iii) To calculate the scores, we assign an equal weight to each reported item. One could potentially assign unequal weights, however, finding an appropriate set of unequal weights is beyond the scope of this work.

The framework presented in this paper can be easily extended to assess the quality of COVID-19 data reporting done by the districts or major cities in India. Another future work is to conduct the same study a few months later and assess the change in the quality of data reporting.

## Data Availability

Links to all data sources and the curated dataset are available in the manuscript.

## 6 Role of the funding source

The funding sources supporting J.Z. had no role in the study design, data collection, data analysis, data interpretation, or writing of this manuscript. All of the authors had full access to all of the data in the study, and had final responsibility for the decision to submit for publication.

## 7 Author contributions

All authors contributed to drafting the manuscript. Scoring metrics were formulated by VV, AG, and JZ. Scoring data was curated and analysed by VV, AG, VS and SAV. JZ supervised the project. All authors reviewed and approved the final manuscript.

## 8 Declaration of interest

All authors declare no competing interests.

## 9 Acknowledgment

The authors would like to thank Professor Steven Goodman of Stanford University for sharing his work on data reporting qualities which inspired this work. The authors would like to thank Minal Patil and an anonymous Stanford student for translating the bulletins released by the states of Gujarat and Andhra Pradesh to English, respectively. We would also like to thank Samuel Joseph, Aravindh Kumar, and Jithin K. Sreedharan for many helpful discussions.

## A Schematic of a good data reporting system

**Figure A.1:**
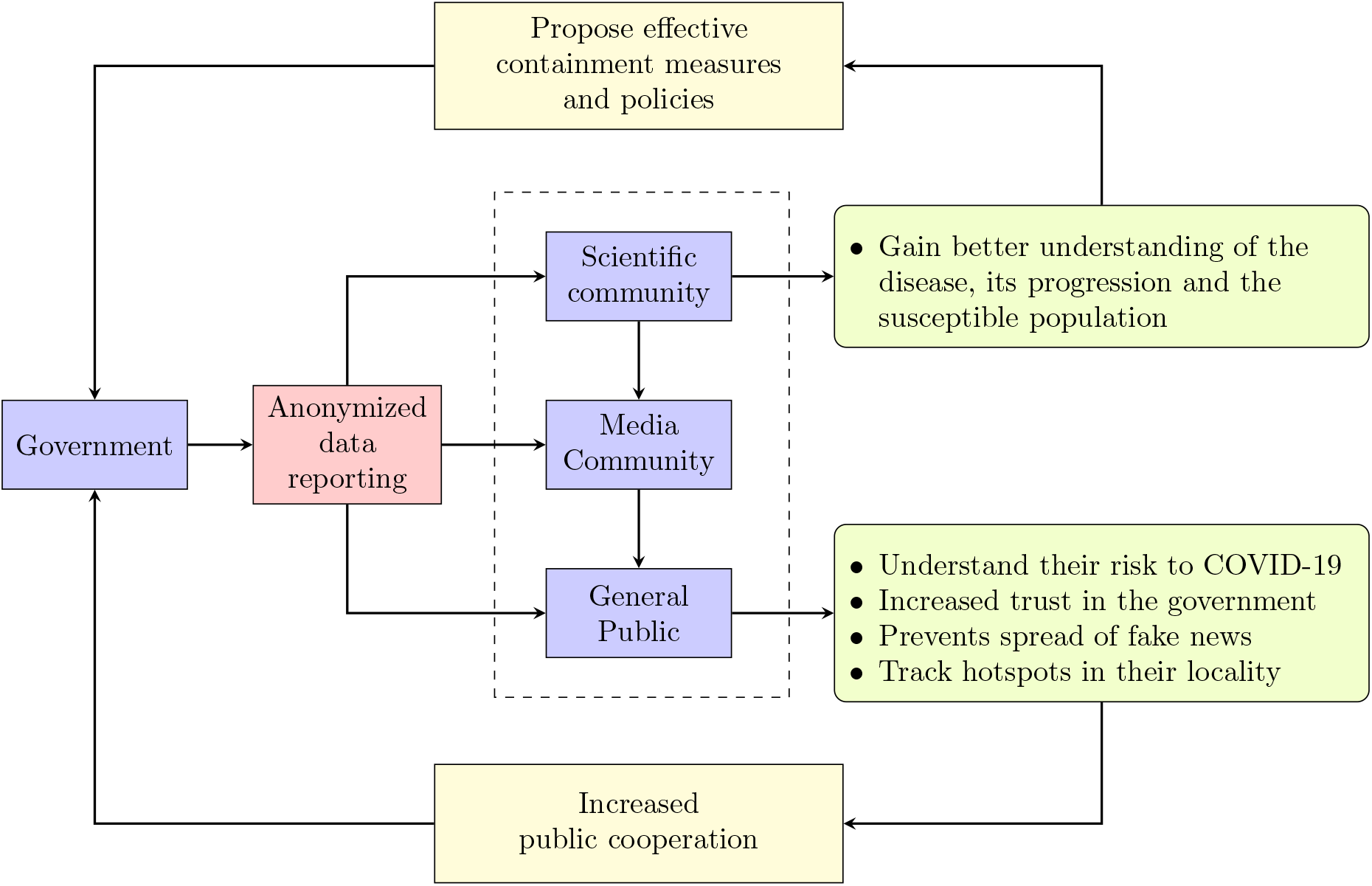
A schematic summarizing the positive outcomes of a good data reporting system.

Figure A.1 shows the schematic of a good data reporting system. As seen from the schematic, data reported by the government is consumed by the general public and the scientific community. The general public consumes data either directly from the government sources or through news media. As shown in Figure A.1, high quality data reporting by the government creates a positive feedback loop that in turn helps the government contain the pandemic better.

## B Template for daily COVID-19 data reporting

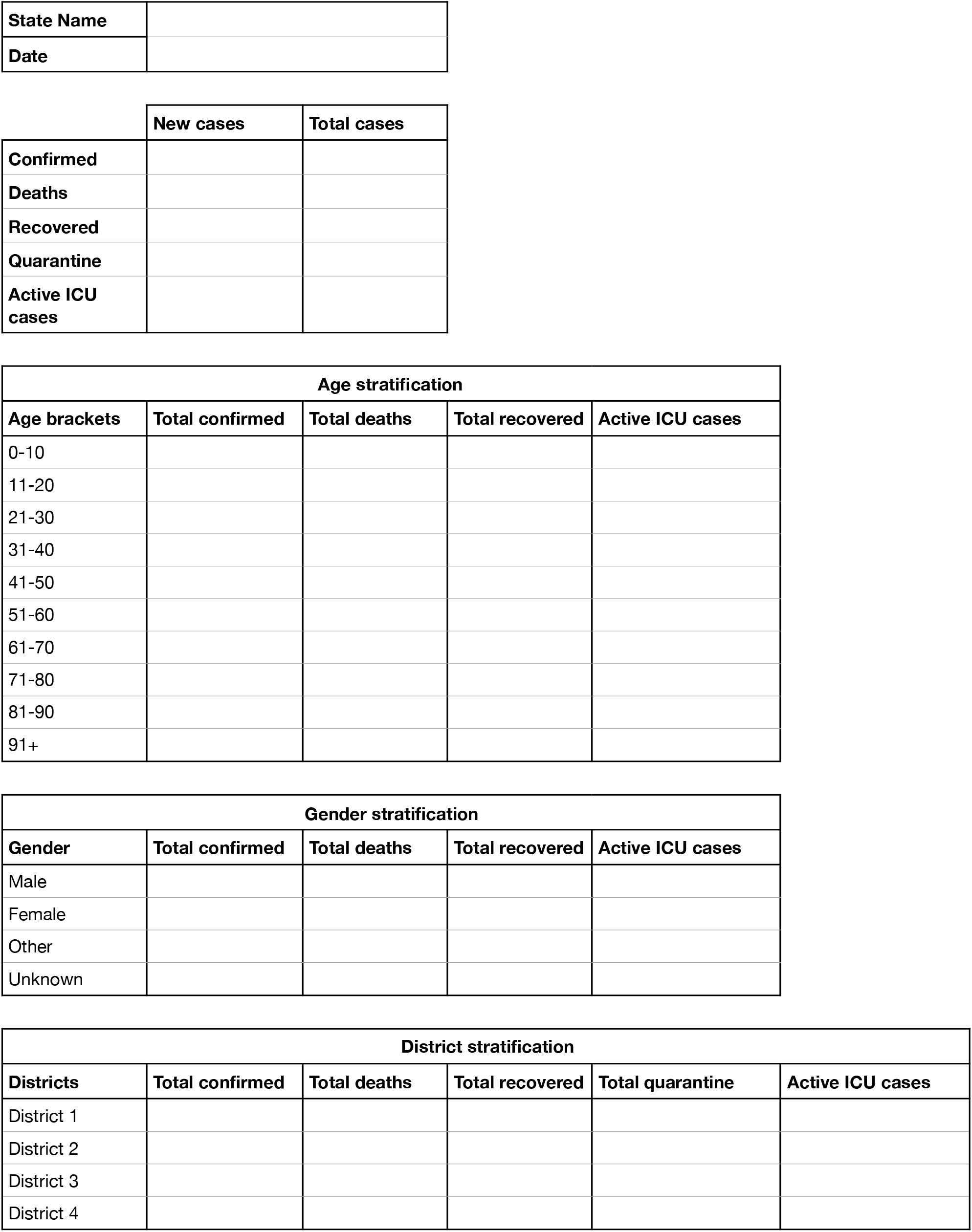
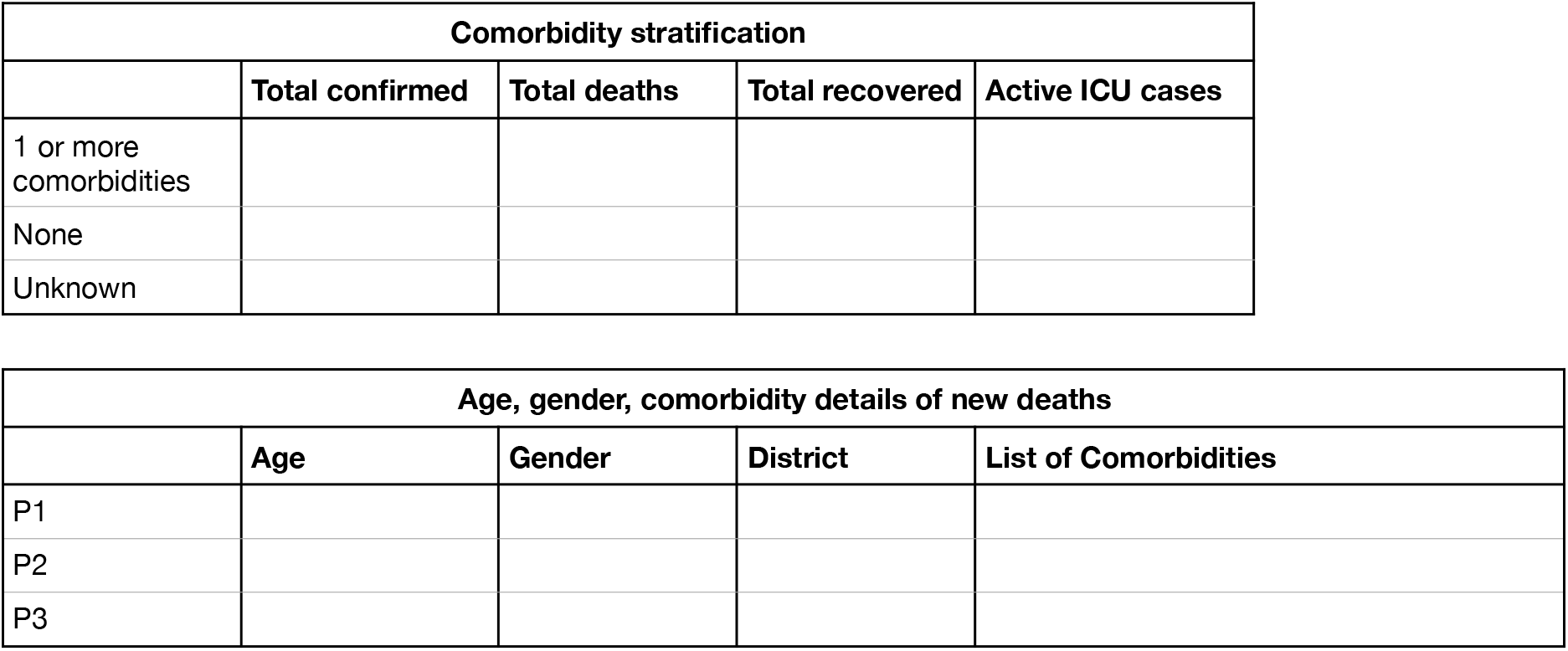

## C Twitter page of covid19india.org

The following are a set of sample questions^v^ asked on the twitter page of covidindia19.org.

a. Request for data at the district level.
  - https://twitter.com/craomumbai/status/1261156322860388352
  - https://twitter.com/HarshuSamnani/status/1246725069646098434
b. Request for death data stratified by age, gender, comorbidities, and districts.
  - https://twitter.com/ramesh_basil/status/1248673419920461825
  - https://twitter.com/nikhilvaishy65/status/1264051034688651265
  - https://twitter.com/UPisute/status/1259667770427359232

## D Details of the scoring metrics

In this section we provide additional information on each scoring metric listed in Table 1 to elucidate what a metric is checking/assessing and its importance in the context of data reporting.

- Total: Total refers to the total as of a given date. This metric checks the availability of total information for each report item. Presence of the total information is assigned a score of 1 and absence a 0 as shown in Table 1. Cumulative data is useful in multiple ways. For example, using total confirmed one can calculate the percentage of population that has been infected and the doubling time of the disease.
- Daily (New): This metric checks the availability of new numbers for each report item on a daily basis. A score of 1 is given for a report item if the new number is available and 0 otherwise.
- Historical data: This refers to the availability of the historical daily data for each report item. Availability of historic data is crucial in determining whether or not the epidemic curve is flattening. A score of 1 is given if historical daily data is available for a report item and 0 otherwise.
- Ease of access: The data is categorized as easily accessible (represented by a 1) if the web page where data is reported is linked from either the state government website or the state health department website.
- Availability in English: Data is marked as available in English (represented by a 1), only if all the items marked as reported in the scoring table are available in English. When the data is reported in English, it is available to a wider audience who speak different regional languages.
- Trend Graphics – Total: This refers to the time-series line chart of the total of a reported item. Date is represented on the horizontal x-axis and total value on the vertical y-axis. The height and slope of the line allows us to see the trends. A score of 1 is given if the trend graphic is present and 0 otherwise.
- Trend Graphics – Daily: This is the same as trend graphics for total, but with daily (new) numbers on the y-axis.
- Stratified by age: This checks if the total for a report item is split into age brackets. For example, a state could stratify the total number of deaths and report the number of deaths in the age groups 0–10, 11–20, …, 81–90 and 91+. This information is helpful to identify and protect the vulnerable population. A score of 1 is given if data stratified by age is available and 0 otherwise.
- Stratified by gender: It refers to the stratification of the total for a report item by gender. Current global data shows that the infection and mortality rate are high among men.^18^ Such inferences would not have been possible if gender stratification of the infected population was not reported. Furthermore, monitoring local trends in the data is useful to improve our understanding of the infection by either confirming or contradicting the global data. A score of 1 is given if data stratified by gender is available and 0 otherwise.
- Stratified by comorbidities: It refers to the stratification of the total for a report item by comorbidity. To keep it simple, if binary stratification (presence/absence of comorbidity) is reported we record a 1 in the scoring metric table. For the case of deaths, a score of 1 is recorded if either of the following information is reported: (i) binary stratification (ii) patient specific details for each death. If both are reported, a score of 2 is assigned.
- Stratified by districts: It refers to the stratification of the total for a report item by districts within the state. For the general public, stratification by districts is way more important than stratification by age, gender and comorbidity. District level information is helpful for the public to understand the effect of the pandemic in their neighborhood, and to cooperate and adhere with government policies and interventions. District level information is also useful for manufacturers of healthcare equipment like personal protective equipment and ventilators, to decide on resource allocation and supply chain logistics. A score of 1 is given if data stratified by districts is available and 0 otherwise.
- Compromise in privacy: This metric checks if any personally identifiable information related to individuals who are quarantined or tested positive for COVID-19 are published online by the government. Examples of identifiable information include name, address, and mobile number. As explained in subsection 2.1, releasing personally identifiable information can have dire consequences. A score of +1 or −1 is entered in the scoring table to indicate “no violation” or “violation” of privacy respectively.

## E Scoring process

During the scoring period, data was curated for each state by filling the scoring metric table shown in Table 1 by following the scoring metrics described in Appendix D. The steps followed to fill the scoring metric table for each state are as follows.

- Authors VV and AG jointly checked the government and health department websites of the state for COVID-19 data on an arbitrary day during the scoring period. Data available on these websites was used to fill the scoring table. If no data was available on either of those websites then a google search^vi^ was done to find other official sources. During the process if any official website was found to contain COVID-19 data, then that was used to fill the scoring table. Social media websites like Twitter and Facebook were excluded. The links to official government websites reporting data are available in Appendix J.
- Two other authors VS and SAV independently verified the entries in the scoring table based on the data reported by the state on another date during the same time period, by following the procedure described above. Any discrepancy/mismatch was noted down for further review by authors VV and AG. The set of states verified by VS and SAV were mutually exclusive.
- For the states that reported data in a regional language that none of the authors could read, external help from a native speaker of that language was obtained to fill the scoring table. There were two states in this category.
- After the scoring period authors VV and AG did a final pass over all the states. During the final pass VV and AG did the following.
  − Addressed the discrepancies/mismatches reported by VS and SAV. If an item was reported when VV and AG filled the table, but not on the day VS or SAV verified, or vice-versa, then that item was marked as unreported.
  − Items in the historical row of the scoring table were marked as reported only if they were reported on all fourteen days during the scoring period.
  − Any item that was not applicable for a state was marked as ‘NA’. For example, (i) stratified by districts is not applicable to Chandigarh, as it doesn’t have any districts; (ii) for states that reported zero deaths until the end of the scoring period, stratified by age, gender, and comorbidities for deaths were marked as ‘NA’. (iii) for a state that doesn’t report any data privacy is marked as ‘NA’.

The curated data is publicly available at this link.

## F Categorical scores

**Figure A.2:**
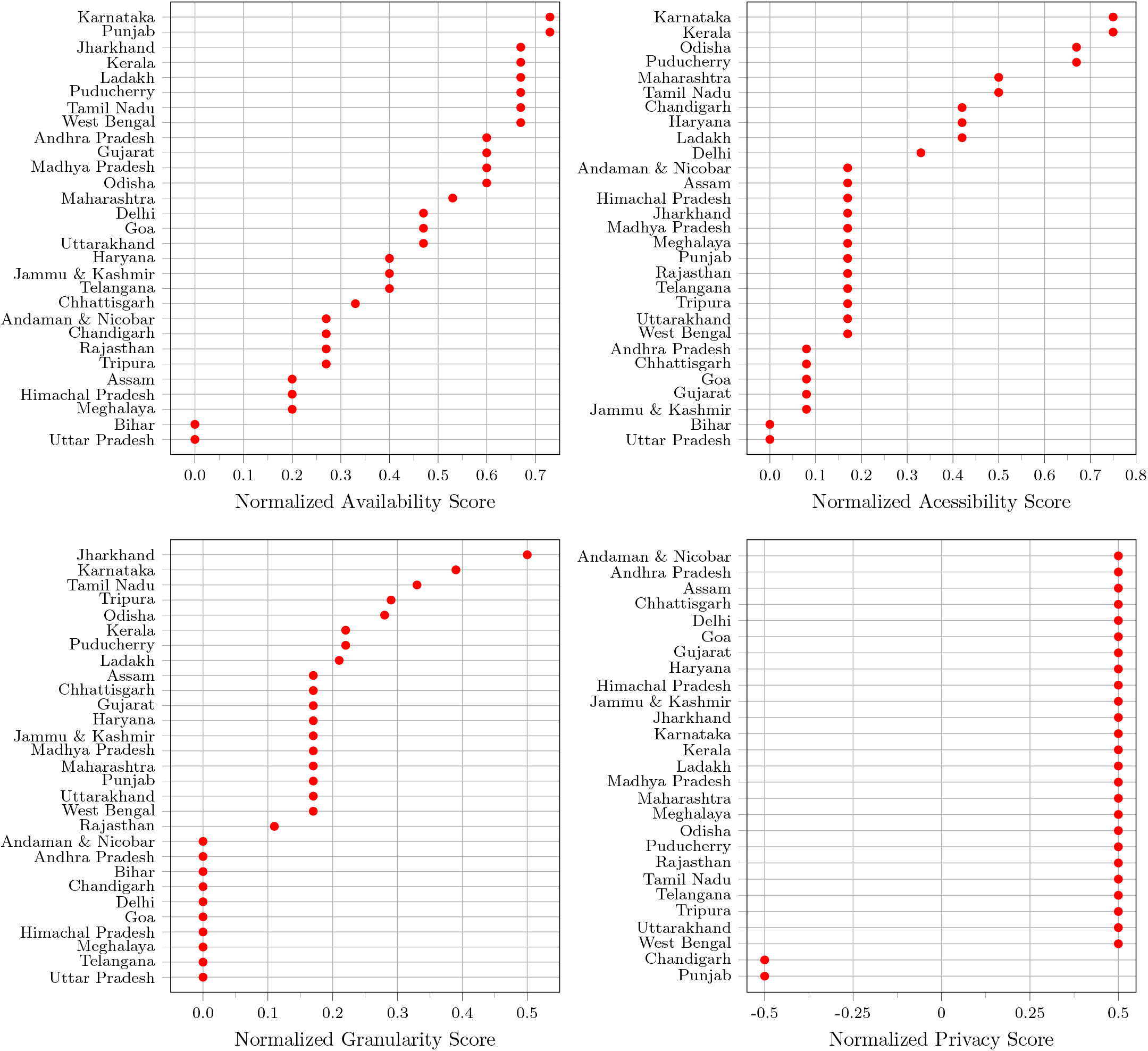
Dot plots showing the normalized availability, accessibility, granularity, and privacy score. Bihar and Uttar Pradesh are not shown in the privacy plot because privacy doesn’t apply to them as they don’t release any data.

CDRS and the normalized categorical scores for the 29 states and union territories are listed in the table below in the alphabetical order.

|  | State /<br>Union Territory | Accessibility<br>Score | Availability<br>Score | Granularity<br>Score | Privacy<br>Score | CDRS |
| --- | --- | --- | --- | --- | --- | --- |
| 1 | Andaman and Nico-<br>bar Islands | 0.17 | 0.27 | 0.00 | 0.5 | 0.17 |
| 2 | Andhra Pradesh | 0.08 | 0.60 | 0.00 | 0.5 | 0.24 |
| 3 | Assam | 0.17 | 0.20 | 0.17 | 0.5 | 0.20 |
| 4 | Bihar | 0.00 | 0.00 | 0.00 | 0.0 | 0.00 |
| 5 | Chandigarh | 0.42 | 0.27 | 0.00 | -0.5 | 0.20 |
| 6 | Chhattisgarh | 0.08 | 0.33 | 0.17 | 0.5 | 0.22 |
| 7 | Delhi | 0.33 | 0.47 | 0.00 | 0.5 | 0.26 |
| 8 | Goa | 0.08 | 0.47 | 0.00 | 0.5 | 0.21 |
| 9 | Gujarat | 0.08 | 0.6 | 0.17 | 0.5 | 0.30 |
| 10 | Haryana | 0.42 | 0.40 | 0.17 | 0.5 | 0.33 |
| 11 | Himachal Pradesh | 0.17 | 0.20 | 0.00 | 0.5 | 0.13 |
| 12 | Jammu and Kashmir | 0.08 | 0.40 | 0.17 | 0.5 | 0.24 |
| 13 | Jharkhand | 0.17 | 0.67 | 0.50 | 0.5 | 0.48 |
| 14 | Karnataka | 0.75 | 0.73 | 0.39 | 0.5 | 0.61 |
| 15 | Kerala | 0.75 | 0.67 | 0.22 | 0.5 | 0.52 |
| 16 | Ladakh | 0.42 | 0.67 | 0.21 | 0.5 | 0.45 |
| 17 | Madhya Pradesh | 0.17 | 0.60 | 0.17 | 0.5 | 0.33 |
| 18 | Maharashtra | 0.50 | 0.53 | 0.17 | 0.5 | 0.39 |
| 19 | Meghalaya | 0.17 | 0.20 | 0.00 | 0.5 | 0.13 |
| 20 | Odisha | 0.67 | 0.60 | 0.28 | 0.5 | 0.50 |
| 21 | Puducherry | 0.67 | 0.67 | 0.22 | 0.5 | 0.50 |
| 22 | Punjab | 0.17 | 0.73 | 0.17 | -0.5 | 0.33 |
| 23 | Rajasthan | 0.17 | 0.27 | 0.11 | 0.5 | 0.20 |
| 24 | Tamil Nadu | 0.50 | 0.67 | 0.33 | 0.5 | 0.50 |
| 25 | Telangana | 0.17 | 0.40 | 0.00 | 0.5 | 0.20 |
| 26 | Tripura | 0.17 | 0.27 | 0.29 | 0.5 | 0.26 |
| 27 | Uttar Pradesh | 0.00 | 0.00 | 0.00 | 0.0 | 0.00 |
| 28 | Uttarakhand | 0.17 | 0.47 | 0.17 | 0.5 | 0.28 |
| 29 | West Bengal | 0.17 | 0.67 | 0.17 | 0.5 | 0.35 |

## G Screenshot from a bulletin published by Jharkhand

**Figure A.3:**
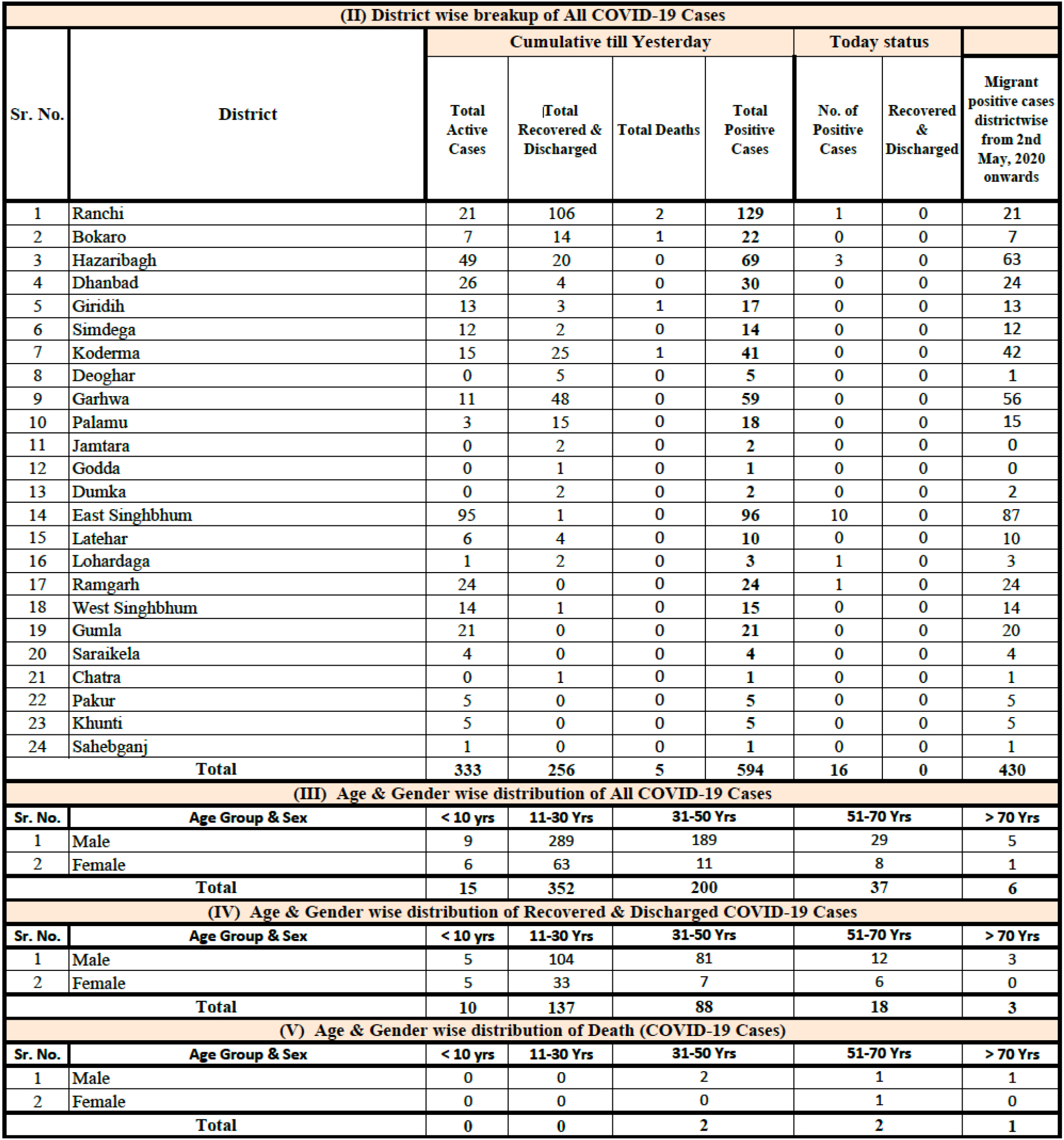
This is a screenshot from the bulletin published by the state of Jharkhand on May 23, 2020. It shows how the state publishes granular data.

## H Additional notes on a few states

- Bihar: The twitter handles https://twitter.com/PIB_Patna/ and https://twitter.com/BiharHealthDept seem to publish some COVID-19 data from Bihar. If we were to consider the data published via twitter, Bihar would get a CDRS between 0 and 0.2. An exact score calculation is not possible without carefully going through all the tweets between 19 May and June 1, 2020.
- Privacy violation in Karnataka: On Mar 25, 2020, a government official from Karnataka released a document containing the details of all persons in a 14-day home quarantine. The document contained residential address (house number, street, district, and pincode) of more than 14k people who were under quarantine. See the original tweet^vii^ and the comments to the tweet at https://twitter.com/bbmpcomm/status/1242726082102456320?lang=en. The release of personally identifiable information like residential address is a clear violation of privacy. Responses to the tweet are a reflection of public concerns about the compromise in privacy. The released document was unavailable during the scoring period of our study — it was probably removed following public concerns. Therefore, we didn’t deduct points for privacy for the state of Karnataka. If we were to deduct points, Karnataka’s alternate CDRS would be 0.57. The alternate score doesn’t change our analysis and conclusions.

## I SDG3-II

The indicators used to calculate SDG3-II are: (1) Maternal mortality ratio, (2) Proportion of institutional deliveries (%), (3) Under-five mortality rate per 1000 live births, (4) Fully immunised children in the age group 0–5 years (%), (5) Total case notification rate of Tuberculosis per 1 lakh population, (6) HIV Incidence per 1000 uninfected population, (7) Currently married women aged 15–49 years who use any modern method of family planning (%), and (8) Total physicians nurses and midwives per 10000 population. For more details on the SDG India Index, check the NITI Aayog website https://niti.gov.in/sdg-india-index-dashboard-2019-20. Note that we assign the same SDG3-II value to both Ladakh and Jammu & Kashmir.

**Figure A.4:**
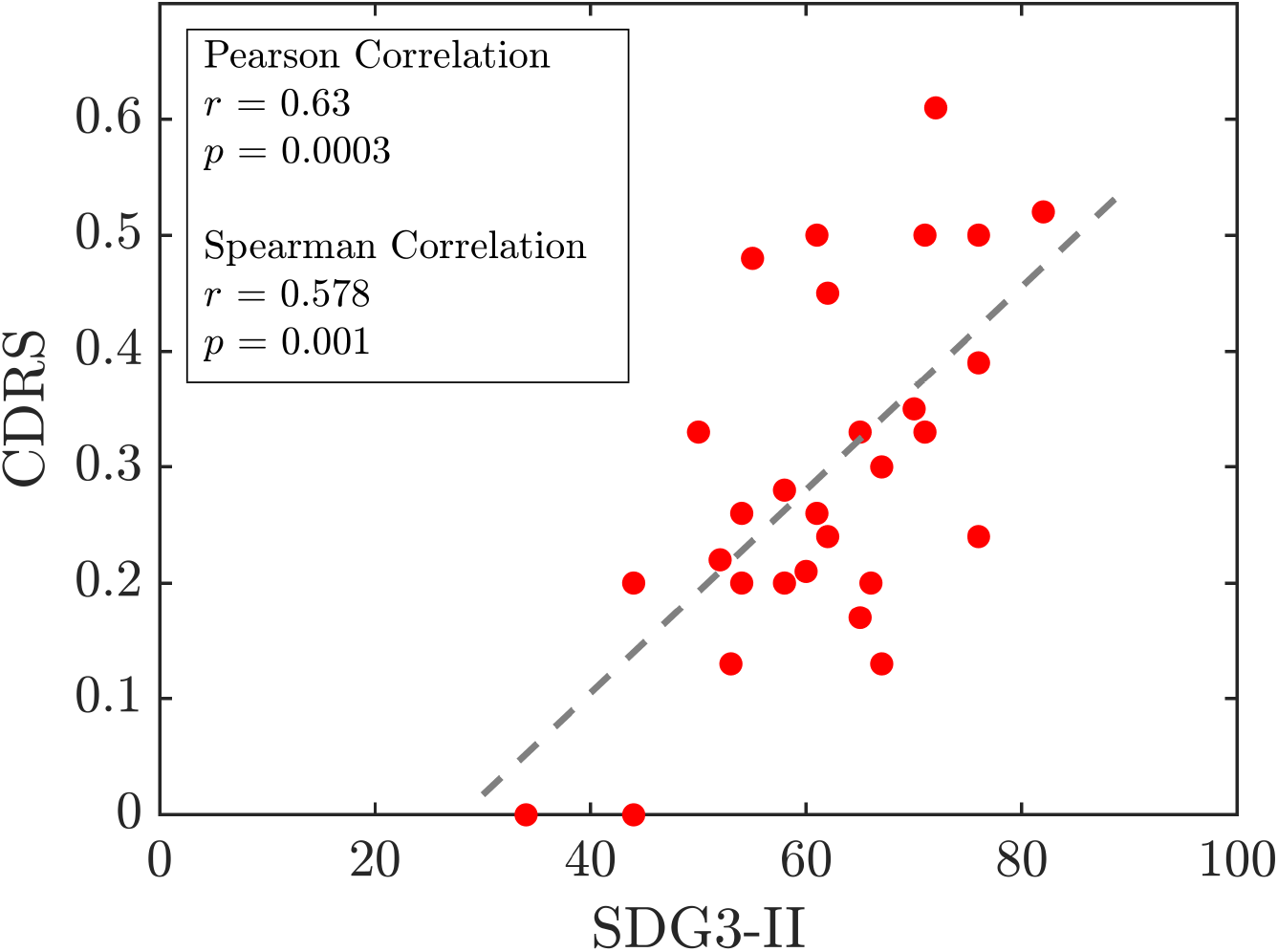
Scatter plot of CDRS versus SDG3-II.

## J Sources for scoring data

|  | State / Union Territory | Data Reporting Websites |
| --- | --- | --- |
| 1 | Andaman and Nicobar Islands | https://dhs.andaman.gov.in/ |
| 2 | Andhra Pradesh | http://hmfw.ap.gov.in/covid_19_dailybulletins.aspx<br>http://hmfw.ap.gov.in/covid_dashboard.aspx |
| 3 | Assam | https://covid19.assam.gov.in/ |
| 4 | Bihar | No sources |
| 5 | Chandigarh | http://chdcovid19.in/ |
| 6 | Chhattisgarh | http://cghealth.nic.in/ehealth/covid19/pages/index.html |
| 7 | Delhi | https://delhifightscorona.in/<br>http://web.delhi.gov.in/wps/wcm/connect/doit_health/Health/Home/Covid19/Bulletin+May+2020 |
| 8 | Goa | https://www.goa.gov.in/covid-19/<br>https://nhm.goa.gov.in/corona-virus-important-links-iec/ |
| 9 | Gujarat | https://gujcovid19.gujarat.gov.in/ |
| 10 | Haryana | http://www.nhmharyana.gov.in/page.aspx?id=208<br>https://gisgmda.maps.arcgis.com/apps/dashboards/5cade394ece3496a9e0c4f168f9536a2 |
| 11 | Himachal Pradesh | http://www.nrhmp.gov.in/ |
| 12 | Jammu and Kashmir | https://www.jkinfonews.com/index.aspx |
| 13 | Jharkhand | https://www.jharkhand.gov.in/Home/Covid19Dashboard |
| 14 | Karnataka | https://covid19.karnataka.gov.in/english |
| 15 | Kerala | https://dashboard.kerala.gov.in/index.php |
| 16 | Ladakh | http://covid.ladakh.gov.in/ |
| 17 | Madhya Pradesh | http://mphealthresponse.nhmmp.gov.in/covid/ |
| 18 | Maharashtra | https://experience.arcgis.com/experience/8167a61f882a4af4b9098e947dfd589f/<br>https://arogya.maharashtra.gov.in/1175/Novel--Corona-Virus |
| 19 | Meghalaya | http://meghalayaonline.gov.in/covid/login.htm |
| 20 | Odisha | https://statedashboard.odisha.gov.in/<br>https://health.odisha.gov.in/ |
| 21 | Puducherry | https://covid19dashboard.py.gov.in/<br>https://covid19.py.gov.in/ |
| 22 | Punjab | https://dronamaps.com/corona.html#/<br>http://pbhealth.gov.in/media-bulletin.htm<br>https://corona.punjab.gov.in/ |
| 23 | Rajasthan | http://www.rajswasthya.nic.in/ |
| 24 | Tamil Nadu | https://stopcorona.tn.gov.in/ |
| 25 | Telangana | http://124.124.103.93/COVID/home.htm |
| 26 | Tripura | https://tripura.gov.in/covid-test<br>https://covid19.tripura.gov.in/<br>https://covid19.tripura.gov.in/Visitor/ViewStatus.aspx |
| 27 | Uttar Pradesh | No sources |
| 28 | Uttarakhand | http://health.uk.gov.in/pages/view/101-covid19-health-bulletin-for-uttarakhand |
| 29 | West Bengal | https://www.wbhealth.gov.in/ |

## K Total confirmed COVID-19 cases as of May 18, 2020

The table below shows the total number of confirmed COVID-19 cases in 29 states and union territories of India as of May 18, 2020. States are sorted in the order of decreasing number of cases. Source: Ministry of Health and Family Welfare.

|  | State / Union Territory | Abbreviation | Total Confirmed COVID-19 Cases |
| --- | --- | --- | --- |
| 1 | Maharashtra | MH | 33053 |
| 2 | Gujarat | GJ | 11379 |
| 3 | Tamil Nadu | TN | 11224 |
| 4 | Delhi | DL | 10054 |
| 5 | Rajasthan | RJ | 5202 |
| 6 | Madhya Pradesh | MP | 4977 |
| 7 | Uttar Pradesh | UP | 4259 |
| 8 | West Bengal | WB | 2677 |
| 9 | Andhra Pradesh | AP | 2407 |
| 10 | Punjab | PB | 1964 |
| 11 | Telangana | TS | 1551 |
| 12 | Bihar | BR | 1262 |
| 13 | Jammu and Kashmir | JK | 1183 |
| 14 | Karnataka | KA | 1147 |
| 15 | Haryana | HR | 910 |
| 16 | Odisha | OD | 828 |
| 17 | Kerala | KL | 601 |
| 18 | Jharkhand | JH | 223 |
| 19 | Chandigarh | CH | 191 |
| 20 | Tripura | TR | 167 |
| 21 | Assam | AS | 101 |
| 22 | Uttarakhand | UK | 92 |
| 23 | Chhattisgarh | CG | 86 |
| 24 | Himachal Pradesh | HP | 80 |
| 25 | Ladakh | LA | 43 |
| 26 | Andaman and Nicobar Islands | AN | 33 |
| 27 | Goa | GA | 29 |
| 28 | Meghalaya | ML | 13 |
| 29 | Puducherry | PY | 13 |

**Figure A.5:**
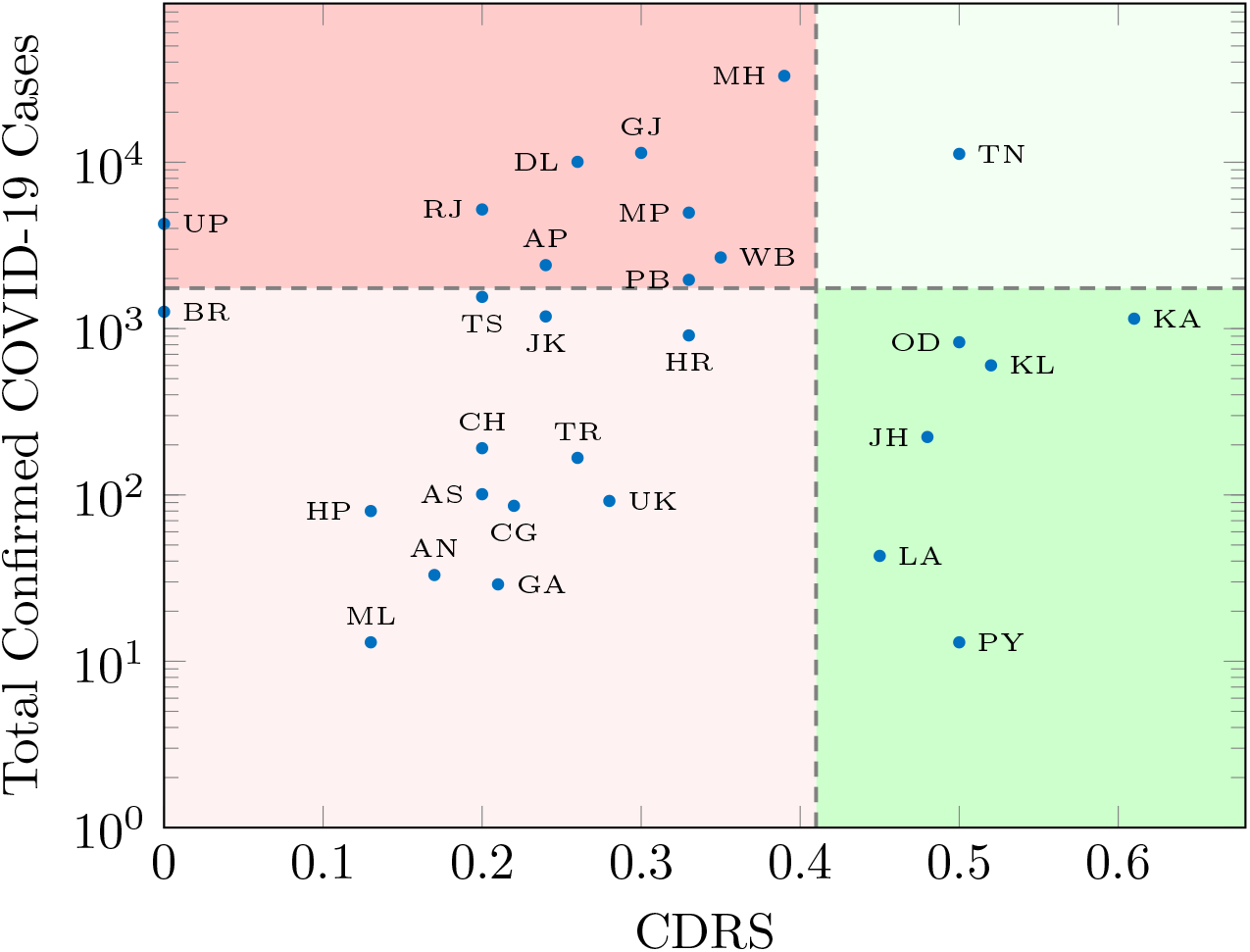
Scatter plot of total confirmed cases versus CDRS. The ten states above the horizontal dashed line contributed to 91% of the total confirmed cases in India as of May 18, 2020. Tamil Nadu is the only state among those 10 with a CDRS in the 75th percentile. The vertical dashed line at 0.41 shows the 75th percentile for CDRS.

## Footnotes

i From here on, unless specified otherwise, the word state refers to a state or union territory in India.

ii Link: https://www.covid19india.org/

iii See Appendix C for a set of sample questions asked on Twitter.

iv Source: Date for the first confirmed case in a state was obtained from Wikipedia.

v These tweets were last accessed on June 28, 2020.

vi Search phrases were of the form “<statename> government covid website” and “<statename> government corona website”.

vii These tweets were last accessed on June 28, 2020.

## Notes

### Competing Interest Statement

The authors have declared no competing interest.

### Author Declarations

The data collected and used in this study were publicly available. Individual consent and ethical approval were not required for the study.

### Summary of Updates

1. Fixed minor typos. 2. Removed research in context section. 3. Author affiliations updated. 4. Edited Fig 2a. caption

